# Diagnostic Prediction with Sequence-of-sets Representation Learning for Clinical Events

**DOI:** 10.1101/2020.08.03.20167569

**Authors:** Tianran Zhang, Muhao Chen, Alex A. T. Bui

**Affiliations:** Department of Bioengineering, UCLA; UCLA Medical & Imaging Informatics (MII); Department of Computer and Information Science, UPenn

**Keywords:** Clinical event sequences, set learning, diagnostic prediction

## Abstract

Electronic health records (EHRs) contain both ordered and unordered chronologies of clinical events that occur during a patient encounter. However, during data preprocessing steps, many predictive models impose a predefined order on unordered clinical events sets (e.g., alphabetical, natural order from the chart, etc.), which is potentially incompatible with the temporal nature of the sequence and predictive task. To address this issue, we propose DPSS, which seeks to capture each patient’s clinical event records as sequences of event sets. For each clinical event set, we assume that the predictive model should be invariant to the order of concurrent events and thus employ a novel permutation sampling mechanism. This paper evaluates the use of this permuted sampling method given different data-driven models for predicting a heart failure (HF) diagnosis in subsequent patient visits. Experimental results using the MIMIC-III dataset show that the permutation sampling mechanism offers improved discriminative power based on the area under the receiver operating curve (AUROC) and precision-recall curve (pr-AUC) metrics as HF diagnosis prediction becomes more robust to different data ordering schemes.

## 1 Introduction

Using the growing amounts of electronic health record (EHR) data, increasing attention has been paid to using data-driven machine learning (ML) methods for a range of classification and predictive tasks, including disease phenotyping and risk stratification [4, 15]. Implicit to these ML-based approaches are a data representation that embodies the temporal nature of such data. One challenge of modeling clinical event data is to learn the representation that aligns with medical knowledge [6, 8, 19], where events (i.e., laboratory results, medications, diagnoses, etc.) can be extracted from time-stamped EHRs and other health-related information, such as claims data. However, many studies modeling such data fail to fully capture the nature of clinical events. For instance, studies modeling claim code sequences only consider temporality between visits, absent of within-visit dynamics [25] that contain essential contextual information. While other approaches utilizing time-stamped EHR events incorporate sequential order within-visit [12, 20], they model a patient’s medical history as a fully ordered event sequence despite the fact that the sequence may contain unordered event sets when multiple events happen concurrently (i.e., sharing the same timestamp). An arbitrary ordering (e.g., random, alphabetical, etc.) is usually imposed on each event set during data preprocessing to establish a “structured” input (e.g., matrices, vectors or tensors) used in different ML models, including contemporary deep learning methods. Consequently, models trained on the corresponding data can be sensitive to the input sequence order as they assume elements from each input sequence to be strictly ordered [30].

The partially-unordered nature of event sequences in the EHR calls for permutation-invariant models: the prediction based on a patient’s medical history should not be affected when the order of concurrent events is changed. In this study, we propose DPSS (Diagnostic Prediction with Sequence-of-Sets), an end-to-end deep learning architecture that incorporates set learning techniques [32] to model event sequences to support downstream diagnostic prediction. DPSS first introduces a permutation sampling technique on each set of concurrent clinical events. A self-attentive gated recurrent unit (GRU) model is then deployed on top of the permutation samples to characterize multiple sets of concurrent events in a patient visit history and correspondingly estimates the risk of specific diseases. To characterize the contextual features of a clinical event, DPSS also pre-trains an embedding model on a collection of unlabeled event sequences. The key contributions of DPSS are threefold: 1) an end-to-end framework modeling clinical temporal event sequences as *sequences of sets* (SoS) for next-visit disease code prediction, with the ability to capture the temporal patterns within each clinical visit; 2) a permutation-invariant prediction mechanism made possible by introducing a permutation sampling technique on SoS; and 3) a demonstration of the utility of a weighted loss function with additional regularization term enforcing permutation-invariant representation of SoS, which further improves the model predictive performance when using permuted sequences. In this way, DPSS is able to represent clinical event data as sequences of sets that are more consistent with the nature of clinical documentation processes.

We evaluate our proposed framework on a binary prediction task for next-visit diagnostic code prediction of heart failure (HF) using laboratory and diagnostic code data from the MIMIC-III dataset [16]. Our experimental results show that approaching clinical event sequence representation from a set learning perspective with permutation sampling more accurately characterizes the underlying disease dynamics and achieves better disease predictive performance. Techniques such as permutation sampling, sequence Laplacian regularization, and self-attention promote permutation invariance and contribute to robustness against different ordering schemes for concurrent events.

## 2 Related Work

### Deep learning on clinical event sequences

Deep learning models, particularly variants of recurrent neural networks (RNN), have achieved some success in modeling sequential data for predictive tasks such as readmission and disease risk [1, 6, 7, 12, 31]. Early efforts in clinical event sequence representation learning focus on constructing low-dimensional representations of medical concepts through word embedding algorithms proposed for natural language processing (NLP) [10, 31]. Key works improved concept embedding by incorporating EHR structures [5, 6, 8, 9] and medical ontologies [29] to capture the inherent relations of medical concepts. More recent methods seek to utilize temporal information, instead of using the indexed ordering, to better characterize chronologies [2, 20, 26, 28]. Still, these aforementioned models mostly assume a fixed temporal order among sequence elements as they serve as inputs, which can cause discrepancies when modeling inputs containing unordered elements.

### Deep set learning

Characterizing heterogeneous feature sets was investigated for applications in point cloud analysis [21, 27, 32, 33] and graph mining [13, 23]. Essentially, a permutation-invariant function is needed for set learning to overcome the limitations of sequence models that are permutation-sensitive [24]. Some of these and other works [24, 27, 32] propose to compress sets of any size into a feature vector using a permutation-invariant pooling operation (e.g., sum/mean/max pooling), although such operations are prone to losing information contained in a feature set [33]. In contrast, permutation sampling-based methods [21, 33] and attention-based methods [18] aim to resolve this issue. For example, Meng et al. [21] specifically use permutation sampling in a hierarchical architecture and concatenation to integrate set element embedding when modeling the structure as a set of sets.

Despite the partially-unordered nature of medical events, only a few studies [25] have been conducted to model clinical event sequences as sequence of sets using a permutation-invariant pooling method. There remains a lack of investigation in the use of permutation sampling strategies on corresponding tasks with EHR-based data, which is the focus of this paper.

## 3 Method

In this section, we first present the design of the proposed framework, DPSS, for next-visit diagnostic code prediction. Fig. 1 illustrates the architecture of DPSS and its three components: 1) a pre-trained lab event embedding layer; 2) an event sequence handler with a permutation sampling mechanism for event sets; and 3) a self-attentive GRU predictor for diagnostic code classification.

**Fig. 1.**
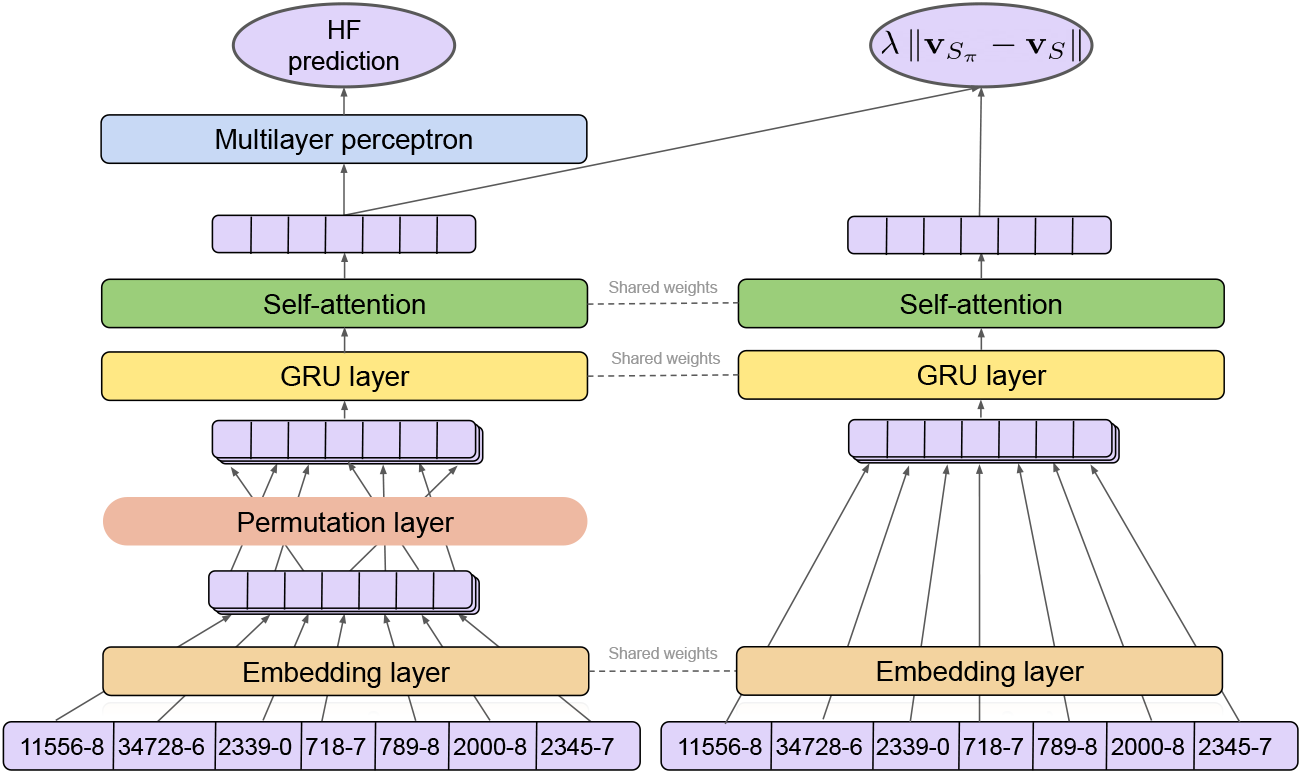
Illustration of DPSS architecture

### 3.1 Preliminary

We use E to denote the vocabulary of lab events, and P to denote the set of patient visit histories. A patient’s visit history in the EHR is defined as a concatenation of lab event sets 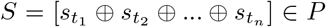, where each set contains lab events with samples collected at the same time *t_k_*, 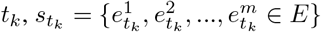. The goal of the diagnostic code prediction task is to provide a regression model to estimate the risk of developing a disease for a patient given the visit history S before the most recent visit. In this case, our goal is to predict codes related to HF.

### 3.2 The DPSS Framework

Our DPSS framework sequentially incorporates three components to characterize and perform prediction on a given patient’s visit history. We first pre-train a lab event embedding model on a large collection of unlabeled historical lab event sequences, which seeks to capture the contextual similarity of lab events. Next, with this pre-trained embedding representing the latent features of each lab event, the permutation sampling process then generates permutations for each event set in the visit history. Lastly, a downstream predictor is trained on the permutation-sampled data, learning to predict the risk for a specific disease while preserving the permutation invariance of concurrent events. Details of each model component is described as follows.

#### Pre-trained lab event embeddings

To encode the non-numerical representations of lab events into numerical representations, we first conduct a pre-training process to obtain an embedding of LOINC codes. We trained a skip-gram language model [22] on a collection of unlabeled lab event sequences with the objective of representing the contextual similarity of lab events in a continuous vector space (obtained by minimizing log likelihood loss):

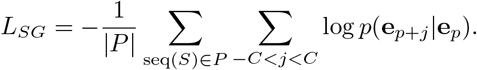

such that seq(*S*) is a temporally-ordered sequence of a visit history 5, and where events in each concurrent set are arbitrarily ordered. Specifically, we extract lab event sequences (from MIMIC-III) as partially-unordered sequences to train the embedding model. *e_p_* is the embedding vector of the *t*-th event *e_t_* ∊ seq(*S*), e*_p_*_+_*_j_* is that of a neighboring event, and *C* is the size of half context.^1^

#### Permutation sampling

Rather than training a decision making model on fixed sequences, the learning objective of DPSS is to make consistent decisions even if such events may be observed in different orders; in our case, this may be dependent on any number of factors as to how an EHR records the data. Inspired by the recent success of deep set learning on point clouds [21, 24, 27, 32], we introduce a permutation sampling strategy for patient visit histories. The principle of this process is to generate event sequences from a given patient’s visit history such that events in a concurrent event set will be randomly ordered in each training epoch, while the sequential order across event sets remain unchanged. In detail, given a set of events *s*, we denote π(*s*) as the set of its permutations. A permutation sample of a visit history S is a sequence 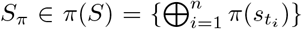 that is obtained by sequentially concatenating a permutation of each concurrent event set in S. Specifically, π(*S*) denotes the universal set of permutation samples for *S*. Based on this sampling strategy, the event sequence encoder introduced next follows an end-to-end learning process for predicting the target diseases, while remaining invariant to the order of concurrent events in a patient visit history.

#### Self-attentive GRU encoder

We use *S*_π_ = [e_1_, e_2_,…, e*_l_*] to denote an input vector sequence corresponding to an embedded lab event sequence after the permutation sampling process of the visit history, *S*. The self-attentive gated recurrent unit (GRU) encoder couples two techniques to represent the embedding representation of the permutation sampled visit history 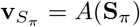.

The GRU is an alternative to a long-short-term memory network (LSTM) [3], which consecutively characterizes sequential information without using separated memory cells [11]. Each unit consists of two types of gates to track the state of the sequence, a reset gate r_p_ and an update gate z_p_. Given the embedding vector *e_p_* of an incoming event, the GRU updates the hidden state 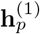 of the sequence as a linear combination of the previous state, 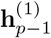, and the candidate state, 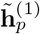 of a new event *e_p_*, calculated as follows:

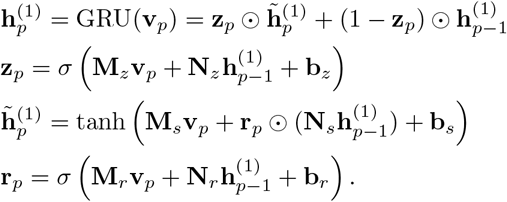

where ⊙ denotes the element-wise multiplication. The update gate z*_p_* balances the information of the previous sequence and the new item, where M_*_ and N_*_ denote different weight matrices, b_*_ are bias vectors, and *σ* is the sigmoid function. The candidate state 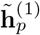 is calculated similarly to those in a traditional recurrent unit, and the reset gate r*_p_* controls how much information of the past sequence contributes to 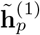.

Atop the GRU hidden states, the self-attention mechanism seeks to learn attention weights that highlight the clinical events that are important to the overall visit history. This mechanism is added to GRU as below:

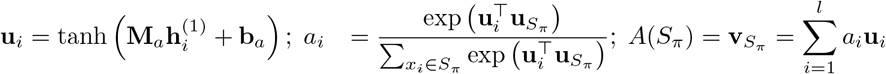

where u*_i_* is the intermediary representation of GRU output 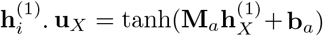 is the intermediary latent representation of the averaged GRU output 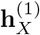 and can be interpreted as a high-level representation of the entire input sequence. By measuring the similarity of each u*_i_* with u*_X_*, the normalized attention weight *a_i_* for 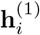 is produced through a softmax function. The final embedding representation 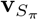 of the visit history is then obtained as the weighted sum of the intermediary representation for each event in the sequence *S_π_*.

#### Learning objective

A multi-layer perceptron (MLP) with sigmoid activation is applied to the previous embedding representation of the visit history, whose output 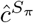 is a scalar that indicates the risk of the target disease. The learning objective is to optimize the loss function defined below.

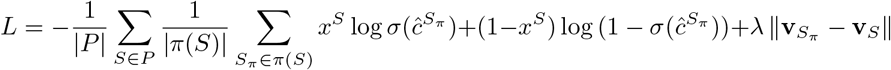

The main loss function uses binary cross-entropy, where *x^S^* ∊ {0, 1} is the training label indicating if the disease code exists in the disease code list from the next patient visit 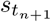. Optimizing for the main loss enforces predictions to be invariant to the input within-set order. The last term of the loss function corresponds to a graph Laplacian regularization term, where *λ* is a small positive coefficient. Notably, this regularization term teaches the self-attentive GRU encoder to generate similar representations for different permutation samples of the same visit history record, and helps differentiate such representations from those of unrelated records in the embedding space. We show below that this regularization mechanism is able to improve the prediction accuracy of the target disease in various experiments.

## 4 Experiments

We hereby evaluate DPSS on the next-visit HF diagnosis prediction task.

### 4.1 Dataset

We evaluated DPSS using data from MIMIC-III [16], a publicly available clinical dataset associated with patients admitted to critical care units of Beth Israel Deaconess Medical Center between 2001 and 2012. MIMIC-III contains records from different sources including demographics, lab results, medications, CPT (Current Procedural Terminology) procedures, and ICD-9 (International Classification of Diseases) diagnostic codes. The within-visit temporal information for diagnostic and procedure codes is not available in MIMIC-III as they are only specific to a patient visit; and while medications are tagged with timestamps, they are recorded with a duration (i.e., start and end times), which poses further challenges on determining the relative ordering between medication and lab events. To simplify our task, we choose to model only lab event sequences as they are less vague with respect to temporal ordering when defined as sequence of sets. Specifically, the timestamp recorded for lab events in MIMIC-III indicates sample acquisition time so a set of lab events with shared timestamps inform patient status at a given time point.

To perform next-visit HF diagnosis prediction, we extracted 7, 235 sequences of abnormal lab events for adult (*age* ≥ 18) patients with at least two hospital admissions from the MIMIC-III dataset by concatenating all abnormal lab events from each visit history. These sequences, each representing a unique patient, are divided into training (75%, 5, 426 patients), validation (12.5%, 904 patients) and test (12.5%, 905 patients) datasets. Based on the existence of the level 3 ICD-9 code representing HF, 428, in the diagnostic codes of the most recent visit, we identified a total of 2, 495 HF cases.We used LOINC codes as the lab event ontology, with 187 unique codes present in our data. During data preprocessing, all eligible event codes for a patient are extracted by patient ID and admission ID matching, sorted by chart time. Concurrent events during the same patient admission are usually imposed with an arbitrary order (e.g., random or alphabetically ordered event codes) when inputted as part of the sequence.

### 4.2 Experimental Configuration

We set the pre-trained skip-gram embedding model on LOINC codes with a context size of 5 and dimensionality of 256. For all reported models, we use the Adam optimizer [17] with a learning rate of 0.001. For each model variant or baseline, we select hyperparameters that lead to the lowest validation loss during training for testing, with the maximum number of epochs set to 100. Training may also be terminated before 100 epochs based on early stopping with a patience of 10 epochs on the validation area under the receiver operator characteristic curve (AUROC) metric. The best combination of GRU layer dimension (candidate values from {64, 128, 256, 512}) and sequence length (candidate values: {128, 256, 512}) is selected based on the AUROC score on the validation set.

We compared the proposed method with the following baseline methods: 1) *GRU*, a single-layer GRU, as defined in Section 3.2; 2) *self-attentive GRU*, a GRU model incorporating the self-attention mechanism; and 3) *Pooling GRU*, following previous work [25, 32], we apply a sum-pooling based or a max-pooling based set function on the set element embedding to acquire a permutation invariant feature aggregation. To show the effects of different model components of DPSS, we also evaluate different variants of DPSS, where we remove the sequence Laplacian regularization or self-attention.

### 4.3 Results

Experiments for baseline models and DPSS are each evaluated on the same holdout test set. We repeated the evaluations 10 times to calculate 95% confidence intervals (CIs) for test AUROC and pr-AUC. Table. 1 summarizes test performance of the baseline models and DPSS.

**Table 1.** Model comparison on next-visit HF risk prediction using MIMIC-III data

| Method | AUROC (95% CI) | pr-AUC (95% CI) |
| --- | --- | --- |
| GRU | 0.7421($\pm 0.00331$) | 0.6133($\pm 0.00564$) |
| Self-attentive GRU | 0.7405($\pm 0.0034$) | 0.6386($\pm 0.0074$) |
| Sum-pooling GRU | 0.7070($\pm 0.00101$) | 0.5839($\pm 0.00173$) |
| Max-pooling GRU | 0.6954($\pm 0.00116$) | 0.5730($\pm 0.00361$) |
| DPSS w/o self-attention&Sequence Laplacian | 0.7741($\pm 0.00277$) | 0.6659($\pm 0.00615$) |
| DPSS w/o Sequence Laplacian | 0.7748($\pm 0.00176$) | 0.6752($\pm 0.00309$) |
| DPSS | **0.7766($\pm 0.00185$)** | **0.6801($\pm 0.00453$)** |

DPSS significantly outperforms the other models in terms of AUROC and pr-AUC metrics. By comparing all of our permutation sampling based model variants with the baseline, we show that the effectiveness of addressing the partially-unordered nature through a permutation sampling mechanism. Specifically, being able to model within-set element interactions, DPSS is shown to be more suitable for modeling lab events as a sequence of sets compared to other permutation-invariant aggregation methods like sum- and max-pooling, with improvements of 9.8% and 11.7% in AUROC, 16.5% and 18.7% in pr-AUC, respectively and relatively. Comparing DPSS variants, we also see that sequentially adding the self-attention mechanism and the sequence Laplacian for permutation-invariant regularization boosted the model’s discriminative power, with greater improvement observed in pr-AUC, which is a metric that considers the model’s ability to cope with imbalanced data [8]. As for the impact of the self-attention mechanism, when added to a basic GRU and DPSS without self-attention and Laplacian loss, the pr-AUC of both models has increased by 4.1% and 1.4%, respectively, while the AUROC metric remained comparable.

We observe that in the raw data of MIMIC-III, concurrent events are ordered randomly in the extracted event sequence. In other data processing scenarios, the event set elements are ordered by the primary key (when applicable) or alphabetically ordered by code strings. The imposed order could lead to bias toward certain data storage methods or a specific coding scheme, which is ultimately irrelevant to the underlying disease. Such inconsistencies may also impair a model’s generalizability when the ordering scheme adopted in training differs from that used during inference. We hypothesized that our set learning framework is able to alleviate the aforementioned bias, as the sequence representation is not restricted to any event set ordering scheme. To test this hypothesis, as our previous experiments are trained and tested on data with random within-set order, we further compared DPSS and the best baseline model against a different event set ordering scheme using test sequences with alphabetically-ordered event sets. These evaluation results are presented in Table. 2.

**Table 2.** Comparison against the best baseline method on the test data with a different ordering scheme (alphabetical) for concurrent events.

| Method | AUROC (95% CI) | pr-AUC (95% CI) |
| --- | --- | --- |
| Self-attentive GRU | 0.7364($\pm 0.00953$) | 0.6214($\pm 0.00878$) |
| DPSS | 0.7755($\pm 0.00305$) | 0.6721($\pm 0.00379$) |

The best baseline model, self-attentive GRU, is trained on set sequences with an imposed arbitrary random order. When tested on alphabetically-ordered set sequences, it suffers from 0.6% decrease in AUROC and 2.7% decrease in pr-AUC. In contrast, DPSS’s performance experienced a smaller decline: 0.1% in AUROC and 1.2% in pr-AUC. The results suggest that DPSS benefited from its permutation sampling mechanism and is more robust against different set ordering schemes.

In summary, the experimental results show that DPSS achieved better performance than the non-permutation sampling-based baseline models on the HF prediction task. The proposed techniques are shown to better capture the clinical events in the visit history according to their partially-unordered nature, hence better supports the downstream decision making.

## 5 Conclusion

We introduce DPSS, a permutation-sampling-based RNN architecture that supports diagnostic prediction with sequence-of-set learning on clinical events. Our proposed method uses a permutation-sampling technique, sequence Laplacian regularization, and self-attention to learn a permutation invariant representation that allows for more accurate prediction for a binary disease prediction task. We also demonstrated the robustness of DPSS against arbitrary set orderings by comparing performance on a test set with an altered set order. For future work, we plan to extend DPSS to jointly model lab event sequences with medication and demographic information. We also seek to better support multi-disease prediction by incorporating structured label representations [14] and leveraging pre-training [34] to improve domain adaptation of DPSS.

## Data Availability

Data involved in this study is publicly available at the MIMIC data site.

https://mimic.physionet.org/

## Footnotes

1 The context of a skip-gram refers to a subsequence of an ordered event sequence seq(*S*) such that the subsequence is of 2*C* + 1 length.

